# Comparison of Capillary Blood Self-Collection using the Tasso-SST Device with Venous Phlebotomy for anti-SARS-CoV-2 Antibody Measurement

**DOI:** 10.1101/2023.03.13.23286935

**Authors:** Elise R. King, Haley E. Garrett, Haley Abernathy, Caitlin A. Cassidy, Cameron R. Cabell, Bonnie E. Shook-Sa, Jonathan J. Juliano, Ross M. Boyce, Allison E. Aiello, Emily J. Ciccone

## Abstract

Measuring seroprevalence over time is a valuable epidemiological tool for improving our understanding of COVID-19 immunity. Due to the large number of collections required for population surveillance as well as concerns about potential infection risk to the collectors, self-collection approaches are being increasingly pursued. To advance this methodology, we collected paired venous and capillary blood samples by routine phlebotomy and Tasso-SST device respectively from 26 participants and measured total immunoglobulin (Ig) and IgG antibodies to the SARS-CoV-2 receptor binding domain (RBD) by enzyme-linked immunosorbent assay (ELISA) on both specimens. Qualitatively, no discrepancies were noted in binary results between Tasso and venipuncture-derived plasma. Furthermore, in vaccinated participants, correlation between Tasso and venous total Ig and IgG specific antibody quantitative levels was high (Total Ig: ρ = 0.72, 95% CI (0.39- 0.90); IgG: ρ = 0.85, 95% CI (0.54, 0.96)). Our results support the use of Tasso at-home collection devices for antibody testing.

## Introduction

Collection of blood samples for seroprevalence studies can be challenging, especially those that are large-scale. In many studies, participants are asked to provide a venous blood sample. This method is logistically challenging, requiring skilled phlebotomists who are frequently in high demand for clinical services, and associated with some discomfort. Additionally, if collection does not occur at participants’ homes, it may add an additional layer of difficulty related to travel to a clinic or study site and can affect participation among socioeconomically disadvantaged individuals. However, household phlebotomy visits pose a risk of COVID-19 transmission between participants and staff, especially as new variants that are more transmissible become more prevalent (1). These concerns with in-person collection methods have highlighted the need for identifying blood self-collection techniques as an alternative (2). Other at-home collection techniques that are used include finger pricking and collection of blood spots. However, collection from the finger can be painful and uncomfortable to self-administer (3). Additionally, serum cannot easily be collected from a blood spot without additional extraction protocols (4). A reliable at-home self-collection method for blood that maximizes study subject comfort and convenience and is easy to process for testing could increase participation and mitigate the transmission of COVID-19 between participants and study staff.

The Tasso-SST at-home collection method is a simple device for serum collection from capillary blood intended for use by lay individuals (5). Its reported ease of use and effectiveness in collecting a viable blood sample makes it ideal for sample collection from a wide range of individuals in a variety of non-laboratory settings, including from those with active COVID-19 infection who are isolating. Furthermore, the cost of the Tasso devices is likely comparable to or less than standard phlebotomy, therefore potentially saving time and money for study staff as well as providing a more comfortable and convenient experience for participants. However, research comparing the Tasso device with phlebotomy for any blood analyte is currently limited (6–8). Given the ongoing COVID-19 pandemic, we assessed the accuracy and precision of antibody measurements of SARS-CoV-2-specific antibodies using the Tasso device as compared to standard venous phlebotomy.

## Materials and Methods

### Study Overview

This analysis was conducted as a sub-study of a longitudinal study that aimed to estimate the incidence of, and risk factors for, SARS-CoV-2 among healthcare personnel (HCP)(9). All HCP participating in the parent longitudinal study were eligible for participation in the Tasso sub-study. The study was approved by the University of North Carolina Institutional Review Board (#20-0942).

### Study Procedures

Subjects enrolled in the parent study were approached regarding Tasso sub-study participation at routine, biweekly, in-person study clinic visits between September 2020 and September 2021. All participants provided informed consent prior to sample collection and could participate in the sub-study at multiple visits.

### Sample Collection

Venous blood was collected in two 8mL Ficoll tubes per parent study protocol (9). Those who consented to the sub-study also simultaneously self-collected a serum sample using the Tasso-SST self-collection kit per the manufacturer’s instructions (5). In brief, to collect the sample, participants cleaned their hands and prepared the skin of the upper lateral arm by rubbing until warm and sterilizing with an alcohol wipe before attaching the device to the skin and collecting capillary blood for 5 minutes (5). Participants followed the written instructions provided in the Tasso kit with verbal assistance from phlebotomists during collection only if needed. The majority of participants used the device independently; rarely, the phlebotomists would physically assist those participants who asked for help with placement of the device on the arm and deployment of the needle. If collection was unsuccessful, the participant was asked to attempt collection again with a second device.

### Sample Processing and Testing

All samples were stored at 4°C within 4 hours and processed within 24 hours of collection (9). Plasma was isolated from Ficoll tubes following centrifugation at 1600 x g for 30 minutes. In a biosafety cabinet, it was then transferred into a 15mL tube without disturbing the hazy layer of peripheral blood mononuclear cells (PBMCs) below. The plasma was then heat-inactivated in a water bath for 30 minutes at 56°C. The heat-inactivated plasma was then centrifuged at 1500 x g for 10 minutes to pellet any remaining red blood cells and protein aggregate. The plasma supernatant was then carefully transferred into sterile 1.5ml O-ring tubes and stored at –80°C until testing (9). Tasso serum self-collection kit capillary blood collection tubes were centrifuged at 1500 x g for 10 minutes. Serum was aliquoted into 0.5 mL microcentrifuge tubes and stored at -80 °C until testing (9).

An enzyme-linked immunosorbent assay (ELISA) was performed on both the plasma and the self-collected Tasso sub-study serum samples in parallel. The test used the recombinant spike protein antigen to measure SARS-CoV-2- specific total immunoglobulin (Ig) and IgG antibodies as previously described (9). To account for plate-to-plate variability, a positive to negative ratio (P/N) was determined by averaging the duplicated OD values of the participant sample and then dividing by the OD value of the negative control. A P/N cut-off of 2.57 for total Ig antibodies and 2.44 for IgG antibodies was determined to achieve 99.3% specificity for Ig total testing and 98.3% for IgG testing with values above this level considered positive and values below considered negative.

### Statistical Analysis

The P/N values from the Tasso serum samples were compared to those from venous plasma samples using Spearman’s Rank Correlation. Confidence intervals were constructed using a bootstrap method with 1000 replications to estimate the 2.5^th^ and 97.5^th^ percentiles of the correlation coefficient. The median paired differences were calculated using Wilcoxon Signed Rank tests. All analyses were conducted for both IgG and Ig outcomes at α = 0.05. Analyses were performed using R version 4.0.5.

## Results

We enrolled 26 unique participants, with three subjects participating twice, for a total of 29 sample pairs. Demographics of the study population are described in Table 1. In general, the sub-study population was similar to the larger parent study cohort with median age of 37 years, minimum age of 21, and maximum age of 65 years. A higher proportion of sub-study participants identified as white and non-Hispanic. Out of all collections, 69% of samples (20/29) were collected after vaccination with an mRNA-based, SARS-CoV-2 vaccine, and 31% (9/29) were collected prior to vaccination.

**Table 1.** Characteristics of Study Participants

|  | Participants in entire cohort<br>(n=213) | Sub-study participants (n=26) |
| --- | --- | --- |
|  | n (%) | n (%) |
| **Age Group (at baseline visit)** |  |  |
| 20-29 | 36 (16.9) | 4 (15.4) |
| 30-39 | 94 (44.1) | 14 (53.8) |
| 40-49 | 56 (26.3) | 6 (23.1) |
| 50-59 | 20 (9.4) | 2 (7.7) |
| 60-69 | 7 (3.3) | 0 (0) |
| **Sex** |  |  |
| Female | 148 (69.5) | 17 (65.4) |
| Male | 65 (30.5) | 9 (34.6) |
| **Race** |  |  |
| White | 152 (71.3) | 23 (88.5) |
| Black or African American | 10 (4.7) | 0 (0) |
| Asian | 11 (5.2) | 1 (3.8) |
| Two or More Races | 11 (5.2) | 1 (3.8) |
| Other/Unknown | 29 (13.6) | 1 (3.8) |
| **Ethnicity** |  |  |
| Not Hispanic or Latinx | 163 (76.5) | 23 (88.5) |
| Hispanic or Latinx | 10 (4.7) | 1 (3.8) |
| Other/Unknown | 40 (18.8) | 2 (7.7) |
| **Health Care Role** |  |  |
| Physician | 77 (36.2) | 11 (42.3) |
| Registered Nurse | 54 (25.4) | 6 (23.1) |
| Ancillary Services | 16 (7.5) | 1 (3.8) |
| Advanced Practice Provider | 13 (6.1) | 1 (3.8) |
| Administrative/Non-Clinical Support Staff | 6 (2.8) | 1 (3.8) |
| Respiratory Therapist | 6 (2.8) | 2 (7.7) |
| Other Therapist | 12 (5.6) | 3 (11.5) |
| Unknown | 22 (10.3) | 0 (0) |
| Other | 7 (3.3) | 1 (3.8) |

Use of the Tasso device failed to collect a sufficient amount of blood in 3.5% (1/29) of collections. The remaining 28 Tasso samples were successfully tested for total Ig antibodies, while 23 of 28 (83%) had sufficient remaining sample for further IgG antibody testing. For comparison, venipuncture was attempted 29 times but was unsuccessful 6.9% (2/29) of the time. All 27 plasma specimens derived from venous blood were tested for Ig and IgG antibodies. Of the 26 participants enrolled, 23 (88.4%) had adequate paired samples to perform total Ig and IgG ELISA assays.

Qualitatively, there was 100% agreement in binary test result (i.e., positive versus negative) between the paired samples for both total Ig and IgG testing among all vaccinated and unvaccinated participants. Quantitatively, the P/N values were higher on average for Tasso-collected serum samples in comparison to venous plasma (Total Ig: median paired difference = 2.57, 95% CI = (1.83, 3.32); IgG: median paired difference = 2.70, 95% CI = (2.71, 34.64)). The total Ig and IgG P/N values from Tasso capillary blood and venous blood (Figure 1, Top Panes) were highly correlated among vaccinated participants (Total Ig: ρ = 0.72, 95% CI = (0.39, 0.90); IgG ρ = 0.85, 95% CI = (0.54, 0.96)). In general, the larger P/N values in vaccinated participants had a smaller percent difference for both IgG and total Ig antibodies P/N values for venous and Tasso collection (Figure 1, Bottom Panes).

**Figure 1.**
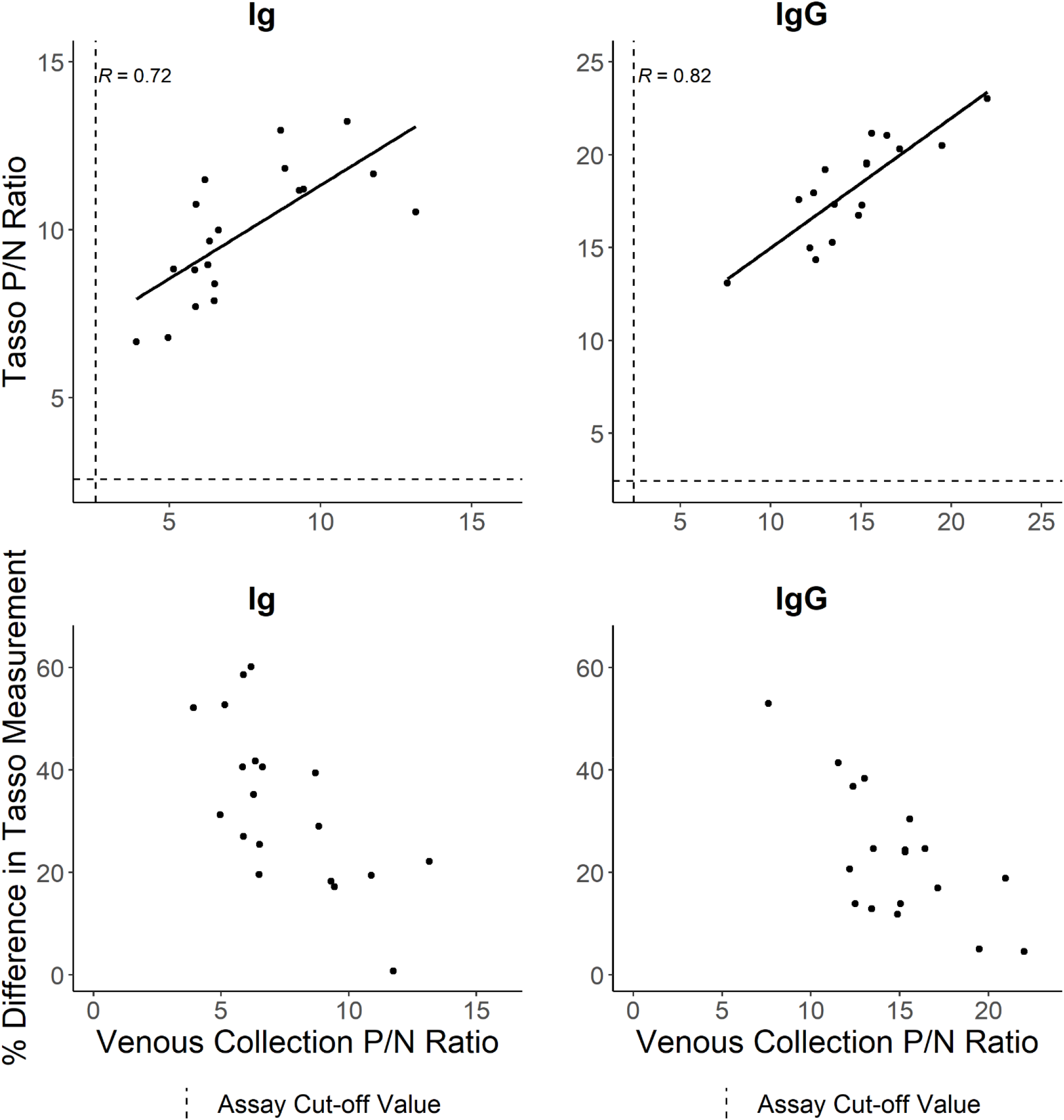
Quantitative total Ig and IgG anti-SARS-CoV-2 antibody measurements from Tasso device self-collection and venous phlebotomy. Top panels show the comparison of the two measurements by Spearman’s Rank Correlation. Bottom panels show the percent difference between the Tasso and venous sample measurements by venous P/N ratio.

## Discussion

In this study, we demonstrated that the Tasso device was a feasible method of collection with adequate sample collected for serological testing for the majority of participants. Additionally, using paired venous blood and Tasso self-collected serum samples, IgG and total Ig measurements were quantitatively similar between the collection methods in vaccinated individuals. Regardless of vaccination status, we found that both plasma from venous blood draws and serum from capillary blood gave the same binary test result. In summary, our findings support that Tasso devices could be used for measuring anti-SARS-CoV-2 antibodies in situations where in-person venous phlebotomy is not feasible or preferable.

We identified three previous studies that have evaluated the performance of the Tasso self-collection kit as compared to venous phlebotomy, and overall, our findings were consistent (6–8). One such study evaluated Tasso’s performance under several conditions: supervised collection of the Tasso device by a healthcare professional and unsupervised at-home self-collection with simulated winter and summer stressors after receipt by the testing laboratory (6). The authors note a good correlation between Tasso and venous phlebotomy collection methods regardless of condition. Our study complements those findings in that we performed simultaneous and monitored, but self-administered, collection of capillary blood using the Tasso device, demonstrating a similarly high correlation with venous blood.

The collection failure and insufficient collection rates of the Tasso device we observed were low and similar to the venous phlebotomy failure rate, although failure with the Tasso was likely less burdensome for participants with no additional needle sticks. One potential way to address Tasso device failure could be to send participants a spare Tasso kit to keep and use if the first collection attempt fails. In addition to 3.5% device failure, 17% of Tasso sample collections did not contain enough serum to conduct more than one ELISA assay, similar to what was seen in another study in which 25% of Tasso kits collected insufficient or no serum (7).

In vaccinated participants, our results show that the P/N values were higher for capillary serum. We suspect this may be due to differences in sample processing. Venous blood was spun, and the plasma was inactivated in a water bath at 56°C for 30 mins. The blood received in the Tasso kit is not heat-inactivated, suggesting that the heat-inactivation may alter the integrity of the antibodies in venous plasma. This would be consistent with previous studies showing that heat inactivation may interfere with the detection of IgG antibody levels (10). However, our data suggest that this does not affect the binary results (positive v. negative) as all paired samples had the same test result.

Our study has several strengths. First, both the venous blood and Tasso samples in our study were kept in a cooler after collection and delivered to the lab within four hours. Therefore, the Tasso samples were not exposed to any adverse conditions or left at room temperature for any extended amount of time as they might when used for at-home collection and mailed prior to testing. This likely reduced the number of external variables that could impact the testing results, such as differences in antibody measurements due to time fluctuations or potential user error of the Tasso device, making our results a good representation of the performance of the Tasso device itself. However, future studies should also address logistical challenges that could impact test results, including those tested in Hendelman, et al (8). Second, previous studies have used the Abbott Alinity nucleocapsid IgG antibody assay (6,7) and the EuroImmun anti-SARS-CoV-2 IgG assay targeting the S1 domain of the spike protein (8). We add to this evidence base by replicating the high correlation between venous blood and Tasso serum when measuring different types of antibodies (both total Ig and IgG antibodies to the receptor binding domain of the spike protein) on another ELISA platform (an assay developed and validated at UNC)(11). The main limitation to our study was the small sample size; it would benefit from replication in the future with a larger subject group. In addition, the low failure rate of Tasso collection could be due to the monitored setting in which the devices were used, and the unsupervised failure rate may be higher than was observed in this study.

## Conclusions

Our findings demonstrate that the Tasso-SST device is an accurate alternative method of blood collection for SARS-CoV-2 serological testing and seroprevalence studies. While this study focused on SARS-CoV-2, it is likely that these results would be generalizable to other analytes and biomarkers that are measured in a similar way. The use of Tasso self-collection devices could decrease the need for clinic collections, reduce the risk of COVID-19 exposure, allow for blood samples to be collected from individuals in all stages of infection, and be more easily implemented in large-scale and low-resource settings.

## Data Availability

All data produced in the present study are available upon reasonable request to the authors

## Acknowledgments

We thank the healthcare personnel participants for their willingness to contribute to advancing our understanding of SARS-CoV-2. We also appreciate the hard work of the phlebotomists who contributed to the sample collection and day-to-day implementation of this sub-study.

## Funding

This project was supported by funds from the following sources:

- 2007 Gillings Gift at the UNC Gillings School of Global Public Health to AEA
- North Carolina Policy□Collaboratory□at the University of North Carolina at Chapel Hill with funding from the North Carolina Coronavirus Relief Fund established and appropriated by the North Carolina General Assembly to RMB
- UNC School of Medicine to RMB
- A generous donation to the UNC School of Medicine by an anonymous donor to RMB
- National Institute of Occupational Safety and Health through Grant Award Number [75D30121P10086 0000HCCK-2021] to AEA and EJC
- National Science Foundation through Grant Award Number NSF 20-4353 to AEA
- National Institutes of Health through Grant Award Number R01 EB0225021 to AEA
- National Center for Advancing Translational Sciences (NCATS), National Institutes of Health, through Grant Award Number UL1TR001111 supported the use of the REDCap database. The content is solely the responsibility of the authors and does not necessarily represent the official views of the NIH.

In addition, EJC was supported by the National Heart, Lung, and Blood Institute through Grant Award Number [5T32HL007106] during the study period, RMB is currently supported by the National Institute of Allergy and Infectious Diseases through Grant Award Number [5K23AI141764], JJJ is currently supported by the National Institute of Allergy and Infectious Diseases through Grant Award Number [5K24AI134990], and BES is supported by the University of North Carolina at Chapel Hill Center for AIDS Research (CFAR), an NIH funded program [P30AI050410].

The funding organizations/sponsors were not involved in the design and conduct of the study; collection, management, analysis, and interpretation of the data; preparation, review, or approval of the manuscript; or the decision to submit the manuscript for publication.

## Conflict of Interest Statement

The authors declare no potential conflicts of interests.

## Notes

### Competing Interest Statement

The authors have declared no competing interest.

### Author Declarations

The Institutional Review Board of the University of North Carolina gave ethical approval for this work.

### Summary of Updates

Corrected typos in the abstract; added caption to Figure 1; added acknowledgments; added funding and conflict of interest statements; corrected references.

